# Charting is Never Exciting: Measuring Physician Time Use in Medical Dramas

**DOI:** 10.1101/2024.03.01.24303412

**Authors:** Katelin C Jackson, Kayla A Miller, Samantha M Hill, Stephanie S. Johnson, Laila A. Reimanis, Eric T Lofgren

## Abstract

**Objectives:** This study aimed to explore the use of medical dramas to train observers when in-person observations or patient contact in clinical settings is impossible. The study also assessed the media’s portrayal of the medical profession and compared time use patterns in medical dramas to previous hospital observational studies.

**Design:** Activity-pattern observational study using Work Observation by Activity Timing software.

**Setting:** The hospital, ICU, and community settings of medical television shows.

**Participants:** The first and last season of the main cast of three medical dramas; Grey’s Anatomy, Scrubs, and ER.

**Main outcome measures:** Inter-rater reliability scores were used to assess how well medical dramas can be used as a training tool for observers. Proportions of time spent on daily medical tasks were compared to other in-person hospital studies.

**Results:** Grey’s Anatomy and Scrubs had excellent Intraclass Correlation Coefficients (ICC) scores for a general medical setting, with Grey’s Anatomy ICC scores for Season 1 0.99 (0.97 to 1.0), and Season 16 0.98 (0.90 to 1.0) and Scrubs ICC scores of 0.91 (0.65 to 0.98) for Season 1 and 0.91 (0.55, 0.99) for Season 8. In contrast, ER had an acceptable ICC score of 0.89 (0.59 to 0.98) for Season 1 and 0.81 (0.35 to 0.96) for Season 15 and might be more suitable for studies exploring activity patterns in the Emergency Department. All had p-values of <0.001.

**Conclusions:** Medical dramas can serve as training tools when clinical observation is limited or impossible, and our methods reflect these shows’ ease of use and flexibility. Additionally, medical dramas can be selected for their similarity to in-person studies. Still, one should be mindful that inaccuracies in the representation of clinical activity patterns are present. However, using medical dramas to train research staff in direct observation is a feasible and reliable method.

## INTRODUCTION

Activity patterns of healthcare professionals (HCP) can have far-reaching implications for the hospital patient-care mission. Prior studies have repeatedly shown that nurse burnout and the work environment have a significant impact on patient outcomes in hospitals [1–3]. The literature on time use and work environment in small to mid-sized community hospitals is limited, and direct empirical estimates are currently lacking. Publications in human health have shown that when directly observing HCPs and recording what activities they perform throughout their day and how often, we can quantitatively assess their time use [4–6]. The Work Observation Method By Activity Timing (WOMBAT) framework has been used to observe the time spent on daily tasks of HCPs [4, 7, 8].

Previous studies have used proximity sensors or direct observations to obtain contact rates which are essential for understanding infectious disease transmission. Sensors have been used to create contact networks in schools [9–12] and healthcare settings [13–15] to understand infectious disease spread. Hospital-based studies that have used sensors measure the effect of HCP number of contacts and the duration of contacts on incidence in patients [13, 15]. However, proximity sensors only record where a healthcare professional (HCP) has been and not what they do. These studies also require a fair amount of infrastructure and, thus, are most commonly conducted in large academic medical centers. Few, if any, sensor-based studies have been conducted in smaller rural or community hospitals.

Direct observation provides more contextual information than sensor-based data; it informs us on what’s actually being done by HCPs throughout the day. These activity patterns can elucidate their patient contact times, who they are in contact with (patient or colleague), and how they spend their overall time. Direct observation has been used to estimate contact parameters to inform analysis on infectious disease spread [13]. While less resource intensive to deploy than sensor-based systems, these methods rely on direct observations, and those making the observations must be adequately trained. Before the pandemic, a study had been planned in a small critical access community hospital in Eastern WA to address the lack of time use estimates in these types of settings. However, the pandemic disrupted the ability to conduct in-person hospital observations due to the infection risk to and from observers; we were forced to find alternative methods to train observers.

### Objectives

In this paper, we explore the use of medical dramas as a means of training observers when in-person observations or patient contact is inaccessible and assessing how the medical profession is portrayed in media. Additionally, we compare time use patterns of observed medical professionals in medical dramas to previously published results from varying hospital settings [4, 7, 8, 16–18].

## METHODS

### Medical Dramas

We selected a variety of medical dramas that represented the daily life of medical professionals, were medically accurate, and were entertaining, namely Grey’s Anatomy, Scrubs, and ER. Mitrokostas, a writer for The Insider — an independent online newspaper, consulted with various doctors and nurses to get their opinions on popular medical shows for their newsletter. Scrubs was deemed the most accurate medical comedy show, with some doctors mentioning “how Scrubs is one of the few medical shows that feels like it was written for doctors” [19]. The show accurately represents what occurs in a hospital on a day-to-day basis and the lives of young resident doctors; it was also the most medically precise.

ER centered around the lives of medical professionals in the emergency department. The doctors and nurses interviewed commented that ER did well in its portrayal of the fast-paced nature of the emergency room and the hectic and exhausting lifestyle of an emergency-care physician [19]. They also mentioned the high medical accuracy of the show, from the proper use of medical jargon to the portrayal of roles and responsibilities of the medical staff and patient representation. However, a doctor commented that the show took creative liberties with the job duties of medical residents, with them doing work they wouldn’t normally do as ER residents [19].

On the other hand, Grey’s Anatomy was rated to be the least medically accurate but highly entertaining among the doctors and nurses interviewed. The most common comments about the inaccuracies of the show were that “interns would never perform surgery without a more experienced physician, hospital superiors being ignored or bypassed, and mistakes being made by residents and them not facing appropriate repercussions” [19]. Another fallacy of the show was that doctors and residents were equally proficient in all types of medicine, such as a “surgical resident delivering babies or involved in complex internal medicine cases” [19].

### Observers And Observations

Observers were undergraduate students in Applied Mathematics and Biology-Pre-Medical who completed their education in Spring 2022 and 2023; they were employed as Research Assistants at Washington State University. They used each medical drama’s first two or three episodes as training and orientation. Observers used the following episodes for training, Grey’s Anatomy: “A Hard Day’s Night”, “The First Cut is the Deepest”, “Winning a Battle, Losing A War”, and “Put on A Happy Face”, Scrubs: “My First Day” and “My Mentor”, ER: “24 hours” and “Day One”. During this time, observers became familiar with the TV show, the cast, the software, and the work definitions. Observers were also allowed to confer with each other on software use and clarify work definitions. Observations were carried out for each medical drama’s first and last seasons at the time of the study. Respectively, these seasons were 1 & 16 for Grey’s Anatomy, 1 & 8 for Scrubs, and 1 & 15 for ER. For each season, a member of the main cast was assigned a number; then, a matching number was assigned to each episode by a random number generator. The selected cast member kept their assigned number for both seasons; however, a new number was assigned if there was a new cast member in a later season.

### Activity Timing

Observers used the Work Observation Method By Activity Timing (WOMBAT) software on the SAMSUNG Galaxy Tab A and iPad Mini tablets. Observers begin recording tasks by selecting what was being done (“What”), whom they were interacting with (“Who”), how they were interacting with an individual (“How”), and where that interaction took place (“Where”) and pressing a button which records the task’s start time. When the observed (participant) begins a new task, more tasks can be selected by pressing the ‘enter’ button.

Additionally, the WOMBAT software allows interruptions and multitasking by pressing the ‘interrupt’ button or the ‘multitask’ button. Television shows have a scripted but structured dialogue, and medical dramas are no different, which is why there were hardly any interruptions being observed. However, when a character had to stop performing an activity and start a new task; then, an interruption was recorded. Observers enter the information for the first task, press the ‘interrupt’ button, and enter the information for the second task. When multitasks occur, the observer must press the ‘add’ button before pressing the ‘enter’ button. If more than one task has ended, individual tasks can be stopped by pressing the ‘end multi’ button. The software requires that at least one task be running at all times. After an observation period (in this case, an episode), the recorded data is uploaded to a cloud server.

### Modifications to the Baseline WOMBAT Definitions

This study primarily used the work definitions from Ballerman et al. 2011. Several categories were added or modified to fit the needs of our study and accommodate the nuances of television. For example, in the “Where” category, “Community”, “ICU,” and “Hospital” were added as options. For the “What” category, “Nothing Happening” was added for periods where dramatic pauses ensured no activities were occurring, and “Offscreen” was added to account for times when the subject of observation was not on screen to be observed. The work definitions are provided in Table 1.

**Table 1.**
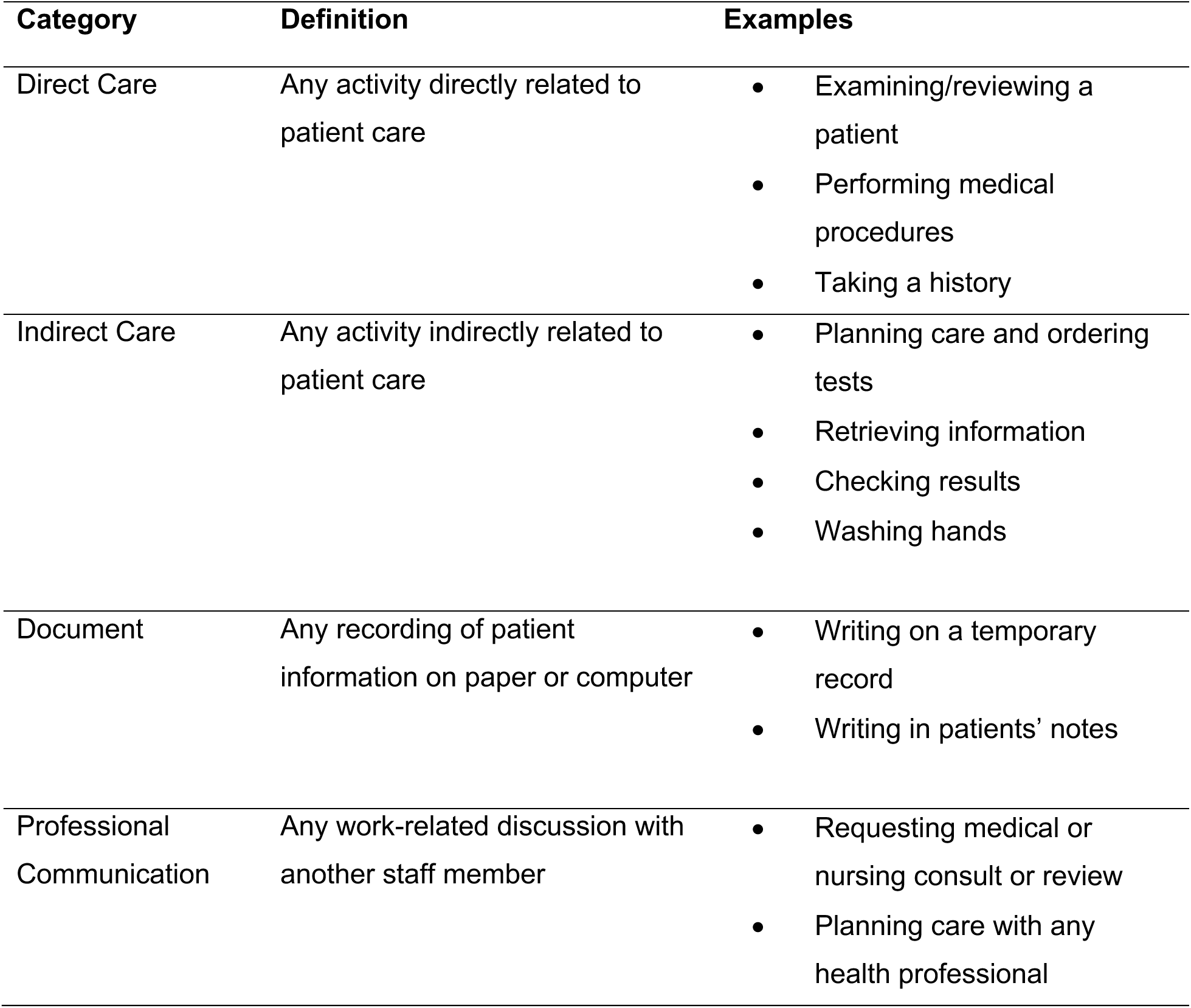
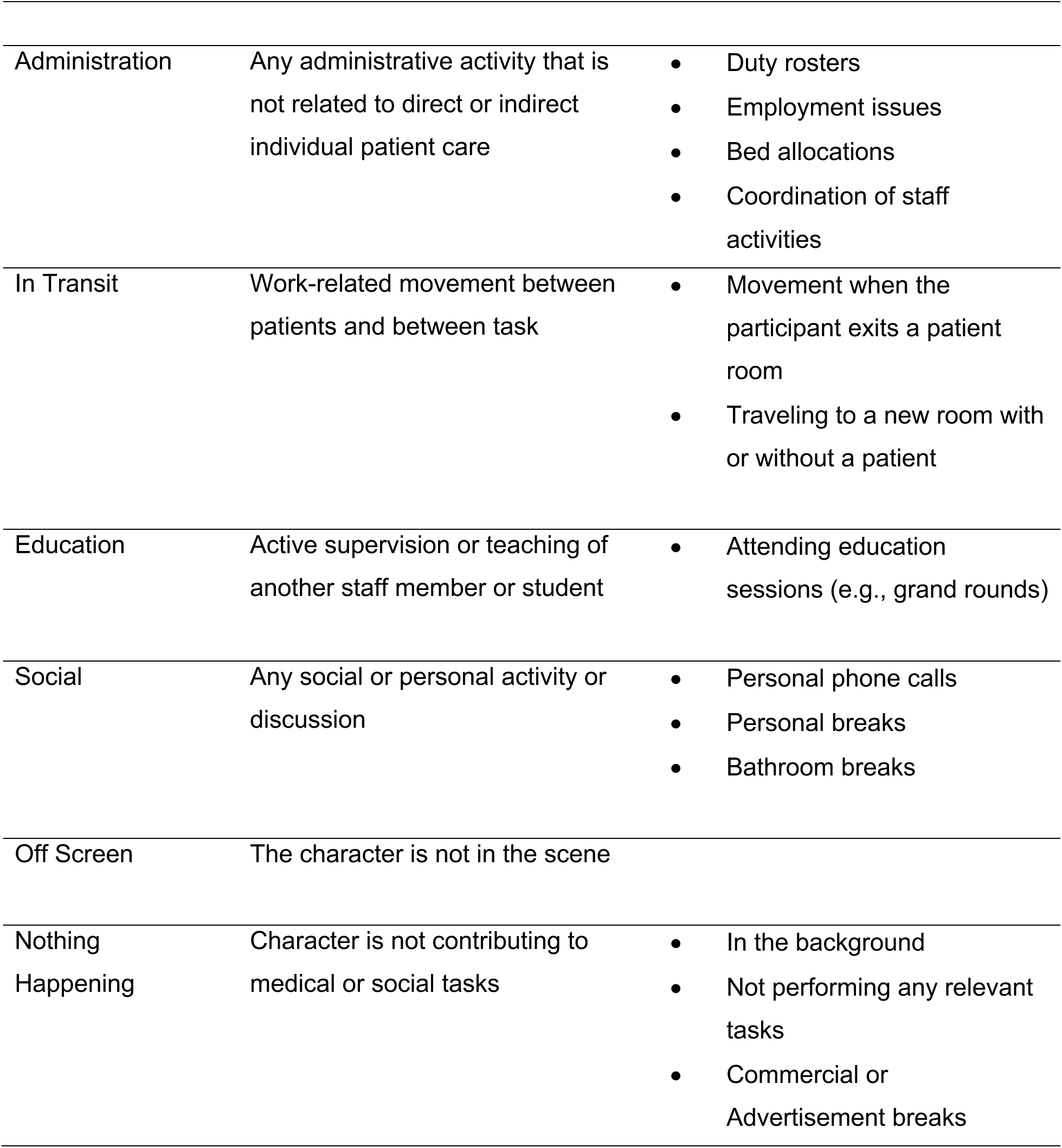
Medical Drama Work Definitions.

| Category | Definition | Examples |
| --- | --- | --- |
| Direct Care | Any activity directly related to patient care | • Examining/reviewing a patient<br>• Performing medical procedures<br>• Taking a history |
| Indirect Care | Any activity indirectly related to patient care | • Planning care and ordering tests<br>• Retrieving information<br>• Checking results<br>• Washing hands |
| Document | Any recording of patient information on paper or computer | • Writing on a temporary record<br>• Writing in patients’ notes |
| Professional Communication | Any work-related discussion with another staff member | • Requesting medical or nursing consult or review<br>• Planning care with any health professional |
| Administration | Any administrative activity that is not related to direct or indirect individual patient care | • Duty rosters<br>• Employment issues<br>• Bed allocations<br>• Coordination of staff activities |
| In Transit | Work-related movement between patients and between task | • Movement when the participant exits a patient room<br>• Traveling to a new room with or without a patient |
| Education | Active supervision or teaching of another staff member or student | • Attending education sessions (e.g., grand rounds) |
| Social | Any social or personal activity or discussion | • Personal phone calls<br>• Personal breaks<br>• Bathroom breaks |
| Off Screen | The character is not in the scene |  |
| Nothing Happening | Character is not contributing to medical or social tasks | • In the background<br>• Not performing any relevant tasks<br>• Commercial or Advertisement breaks |

### Statistical Analysis

A preliminary data cleaning was performed in Excel, then imported into R statistical software (v4.2.1) for further analysis on the proportion of time spent on each categorical task for both the first and last season of all medical dramas [20–22]. Descriptive statistics were used to determine the total observation time, total time per task, and frequency of tasks. The proportion of time spent on tasks and 95% confidence intervals were calculated.

For medical dramas to be an effective training tool, observers should reach the same conclusion. As such, we also considered inter-rater reliability. We used the Intraclass Correlation Coefficient (ICC) via the ‘psych’ R package (v 2.2.9) [23] to measure the strength of inter-rater agreement for continuous variables. A high ICC, close to one, has more agreement between observers. However, an ICC close to zero has less agreement between observers [24], [25]. Values between 0.8 (80%) and 0.89 (89%) are considered good inter-rater reliability, while 0.9 (90%) and 1 (100%) are considered excellent inter-rater reliability [25]. A Chi-square independence test was used to determine the association between each medical drama’s first and last seasons, gender, and the tasks being performed. A Fishers Exact test was used when appropriate due to small cell counts.

### Human Subjects

This research was determined not to constitute human subjects research by the Washington State University Institutional Review Board.

### Patient and Public Involvement

Patients and the public were not involved in any way in this research study.

## RESULTS

We observed 5883 separate tasks across the three chosen medical dramas—the main characters were observed for a total time of 38.72 hours.

### Grey’s Anatomy

Season 1 of Grey’s Anatomy had higher proportions of time spent (29.1%, 95%CI: 28.4% to 29.9%) on direct care tasks. When compared to the latest season, which revealed a decrease in medical tasks but had higher proportions of time spent on social tasks. Professional Communication varied slightly but was the most consistent medical task completed for the seasons compared to other activities (Figure 1). Compared to existing observations from Ballerman et al. 2011, Westbrook et al. 2008, and Westbrook et al. 2010, both seasons dramatically overrepresent physicians’ time directly caring for patients and underrepresenting their administrative duties. Overall, Grey’s Anatomy had more females spending more time on tasks than males, with similar activity patterns between seasons 1 and 16 (Figure 2). As with our previous results for Grey’s Anatomy, professional communication was higher for both seasons, (Season 1: 29.3%, 95%CI: 28.5% to 30%) (Season 16: 29.1%, 95%CI: 28.4% to 29.7%), followed by direct care tasks (Season 1: 23.5%, 95%CI: 22.8 % to 24.2%) (Season 16: 15.8%, 95%CI:15.3% to 16.3%). Moreover, the time spent on social tasks increased among males in season 16 from 1.2% (95%CI: 1.0% to 1.4%) to 11.1% (95%CI: 10.6% to 11.5%), while direct care tasks decreased between females to 15.8% (95%CI: 15.2 to 16.3).

**Figure 1.**
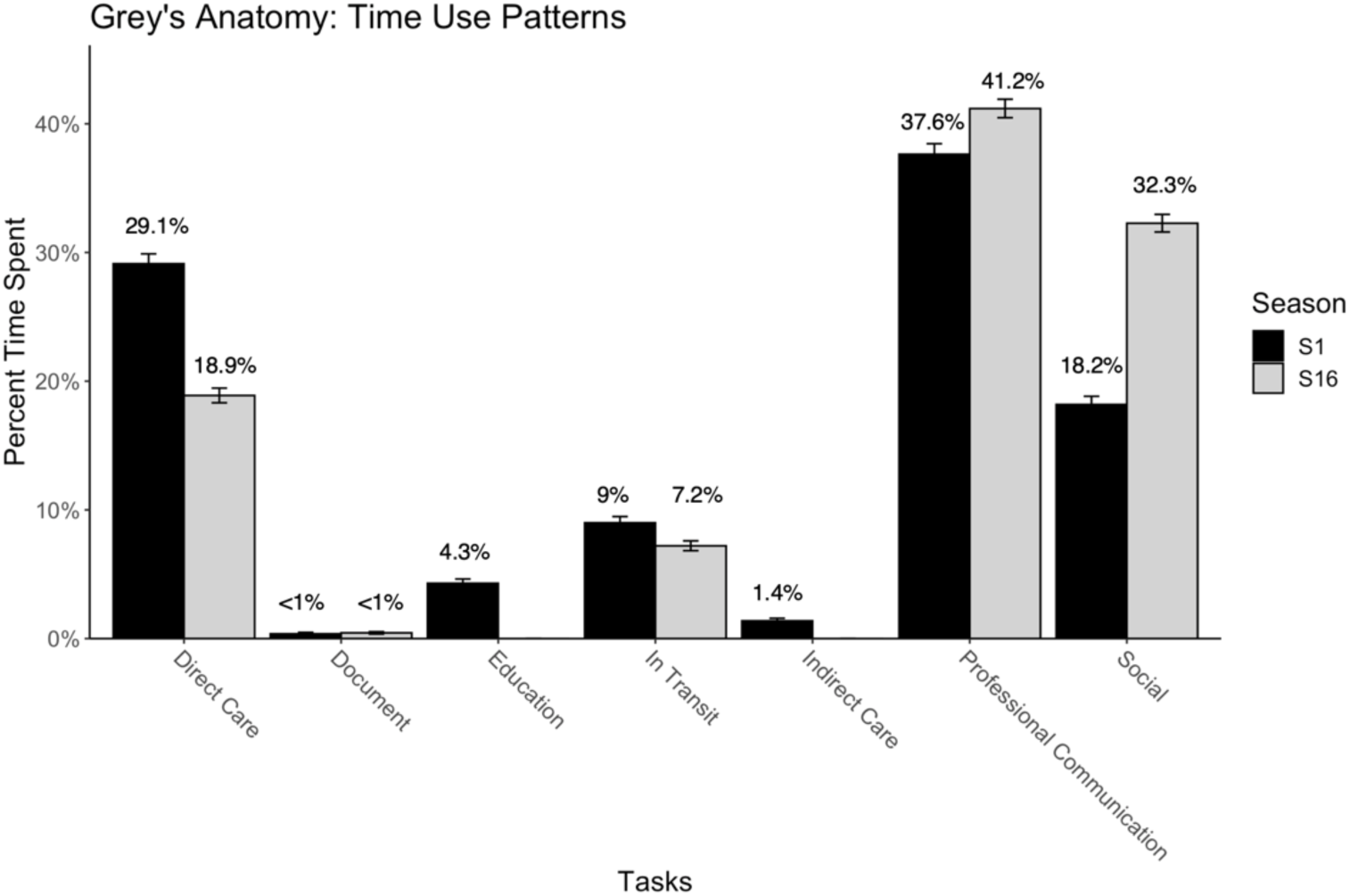
Percentage of time main characters spent on tasks during Grey’s Anatomy’s season 1 and season 16. Error bars are 95% confidence intervals.

**Figure 2.**
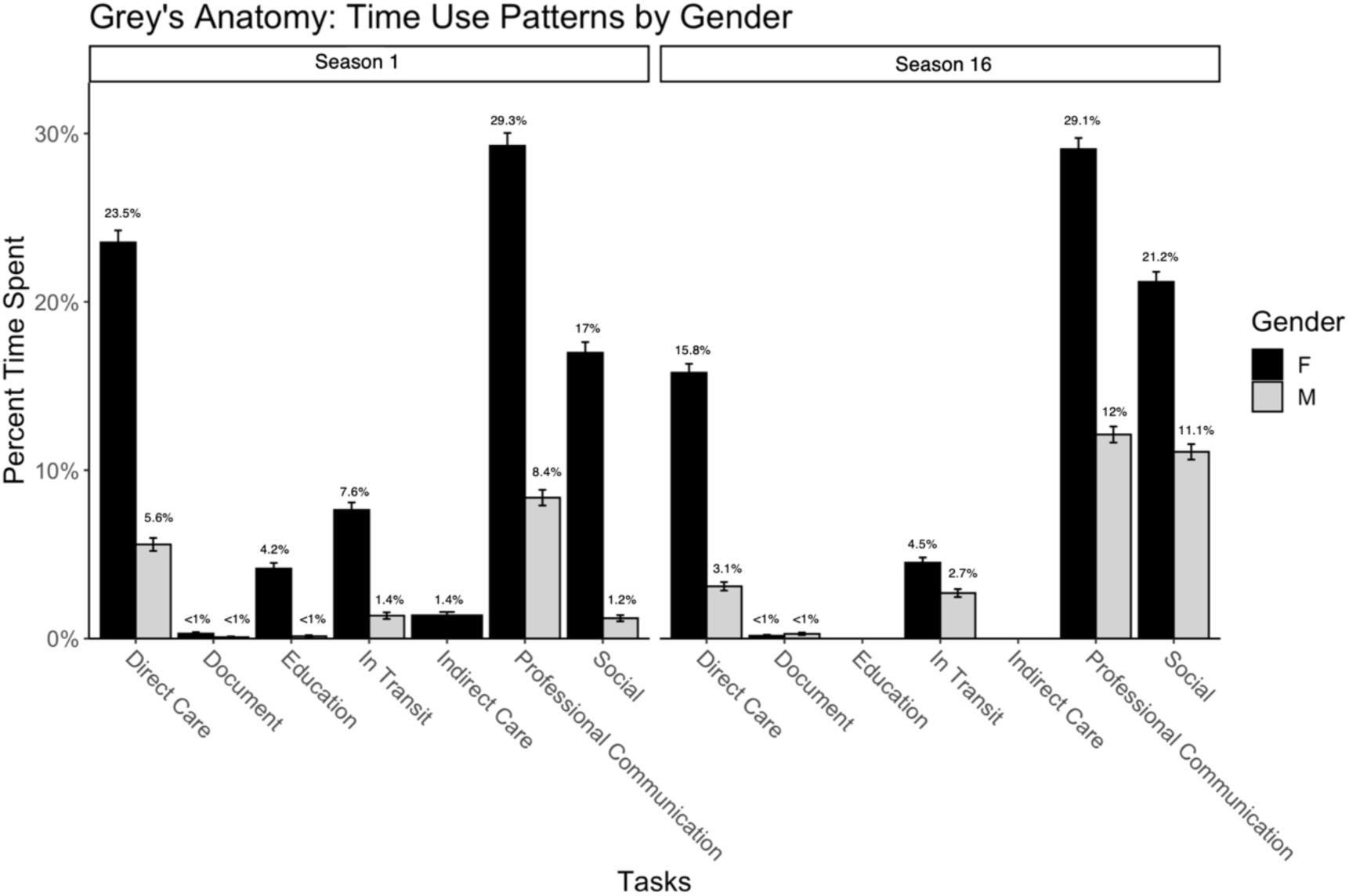
Percentage of time spent on tasks by gender during Grey’s Anatomy’s season 1 and season 16. Error bars are 95% confidence intervals.

A Fisher’s Exact test was performed to determine the relationship between Grey’s Anatomy seasons and the tasks performed. The relationship between these variables was significant, *p* < 0.001. Gender was also considered; we found a significant association between gender and tasks when conducting a chi-square independence test, X^2^ (6, *N=* 1117) = 99.48, *p* < 0.001. The ICC was computed to assess the agreement between two observers in observing medical residents’ and physicians’ time use patterns in Grey’s Anatomy. Critically, for a training tool, inter-rater reliability was excellent, with an ICC of 0.99 and 0.98 for seasons 1 and 16, respectively.

### Scrubs

Scrubs spent more time on social tasks (56.9%, 95%CI: 56.1% to 57.7%) in Season 8 than in the other medical dramas. Hardly any time was spent on medical tasks and administrative duties in Season 1 and Season 8 (Figure 3). But compared to Walter et al. 2014 and others [4, 18], Scrubs is comparable with the realities of medical professionals’ daily duties with direct care tasks (Season 1: 8.7%, 95%CI: 7.8% to 8.5%) (Season 8: 3.5%, 95%CI: 3.3% to 3.8%). Although indirect care tasks (Season 1: 4.9%, 95%CI: 4.7% to 5.2%) (Season 8: 2.6%, 95%CI: 2.4% to 2.9%) were only similar to Ballerman et al. 2011 findings. However, administrative tasks were underrepresented. Professional communication and social tasks had similar proportions of time spent in season 1, with 39.6% (95%CI: 39.0% to 40.2%) and 36.4% (95%CI: 35.8% to 37.0%), respectively. Males contributed more to social tasks in both seasons than females at 29.6% (95%CI: 29.1% to 30.2%) and 48.2% (95%CI: 47.4% to 49.0%) (Figure 4). Direct care tasks were about equal in season 8, but males spent more time on direct care tasks than females in season 1. Time spent on professional communication decreased among males from 34% (95%CI: 33.4% to 34.6%) to 19.3% (95%CI: 18.7% to 19.9%). And there weren’t any drastic changes in female activity patterns for either season.

**Figure 3.**
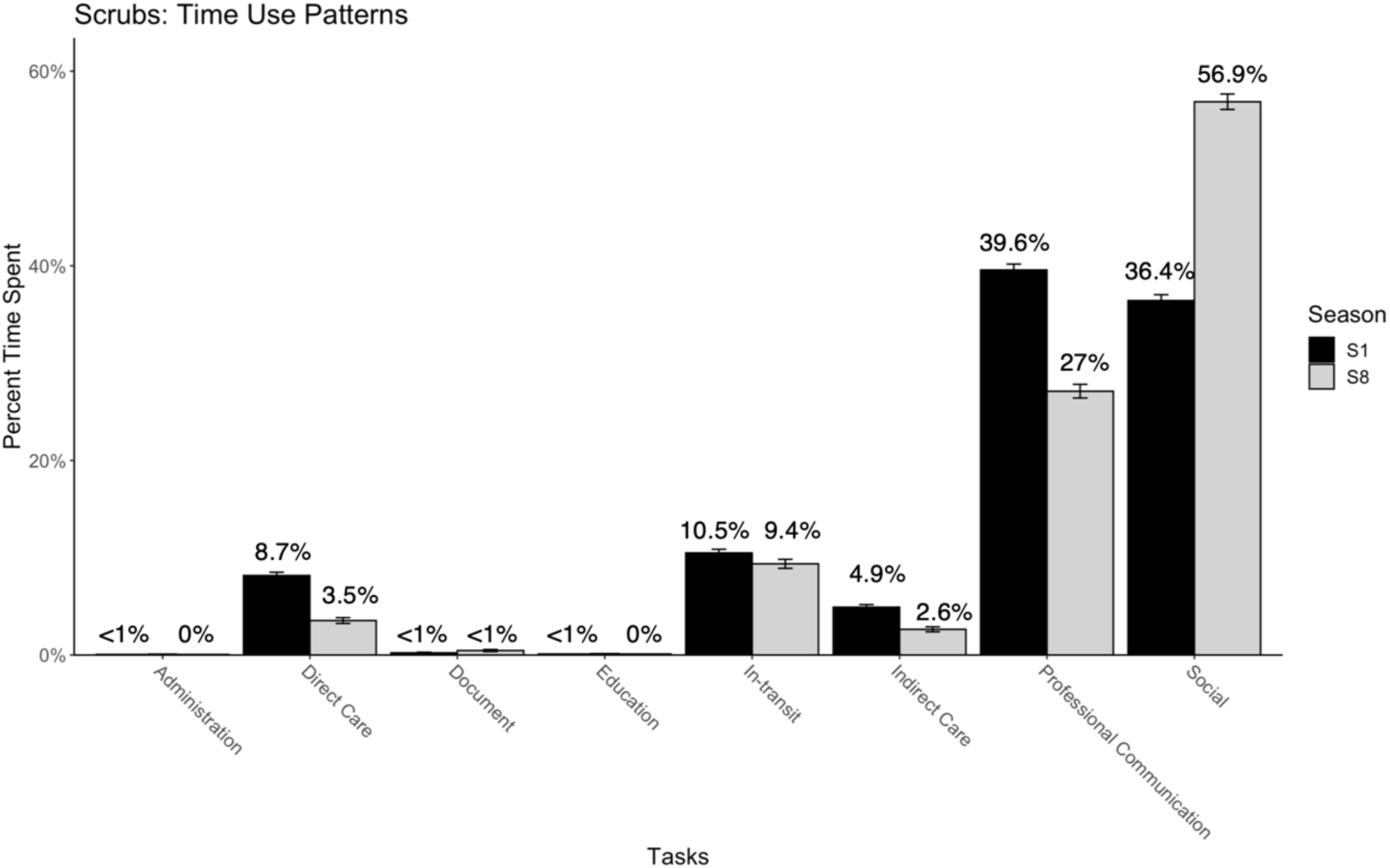
Percentage of time main characters spent on tasks during Scrubs season 1 and season 8. Error bars are 95% confidence intervals.

**Figure 4.**
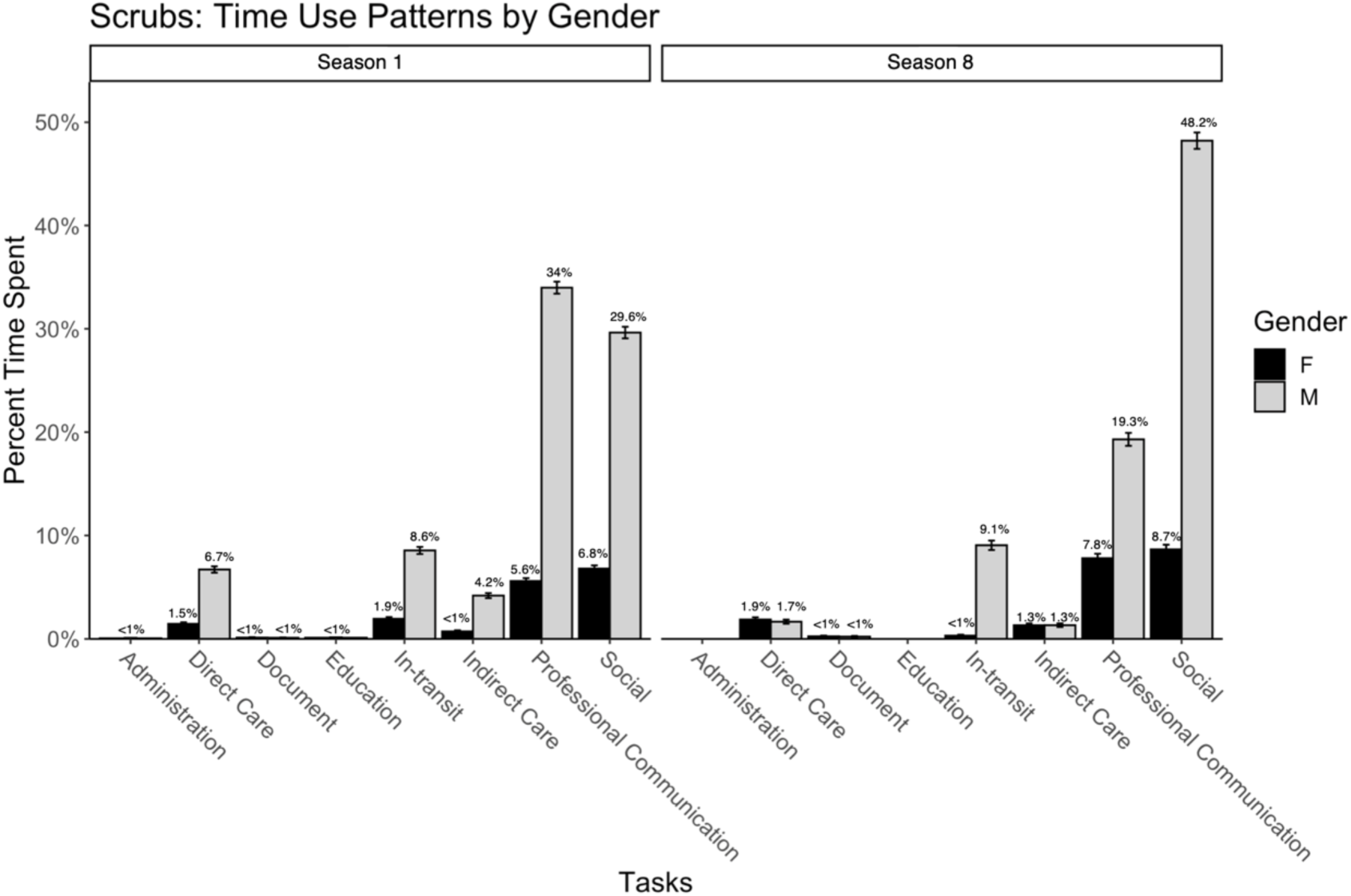
Percentage of time spent on tasks by gender during Scrubs season 1 and season 8. Error bars are 95% confidence intervals.

A chi-square test of independence showed a significant relationship between Scrubs seasons and tasks performed, X^2^ (7, *N=* 1797) = 67.21, *p* < 0.001, and showed a significant association between gender and time use patterns, X^2^ (7, *N=* 1797) = 17.041, *p* = 0.017. The ICC was computed to assess the agreement between two observers in observing medical residents’ and physicians’ time use patterns in Scrubs. The inter-rater reliability was excellent, with an ICC of 0.91 for seasons 1 and 8.

### ER

ER had the highest percent time spent on direct care tasks (Season 1: 36.2%, 95%CI: 35.7% to 36.7%) (Season 15: 43.8%, 95%CI: 43.3% to 44.4%) in both Season 1 and Season 15 than any other observed medical drama. Characters were consistent with performing professional communication (Season 1: 28.3%, 95%CI: 27.8% to 28.7%) (Season 15: 29.3%, 95%CI: 28.8% to 29.8%) and social tasks (Season 1: 17.4%, 95%CI: 7.8% to 8.5%) (Season 15: 17.2%, 95%CI: 17.0% to 17.8%) (Figure 5). Interestingly, ER had the lowest proportion of time spent on social tasks but the highest on in-transit duties (Season 1: 12.4%, 95%CI: 12.0% to 12.7%) (Season 15: 7.6%, 95%CI: 7.3% to 7.9%). Most studies have focused on general practice or ICU physicians’ activity patterns. Still, ER had similar proportions of time spent on professional communication but higher in direct care tasks (Season 1: 36.2%, 95%CI: 35.7% to 36.7%) (Season 15: 43.8%, 95%CI:43.3% to 44.4%) compared to other studies interested in the activity patterns of the Emergency Department [4, 16]. Throughout both seasons, females and males showed the same activity patterns between direct care, professional communication, and social tasks, with males contributing to these tasks most of the time (Figure 6).

**Figure 5.**
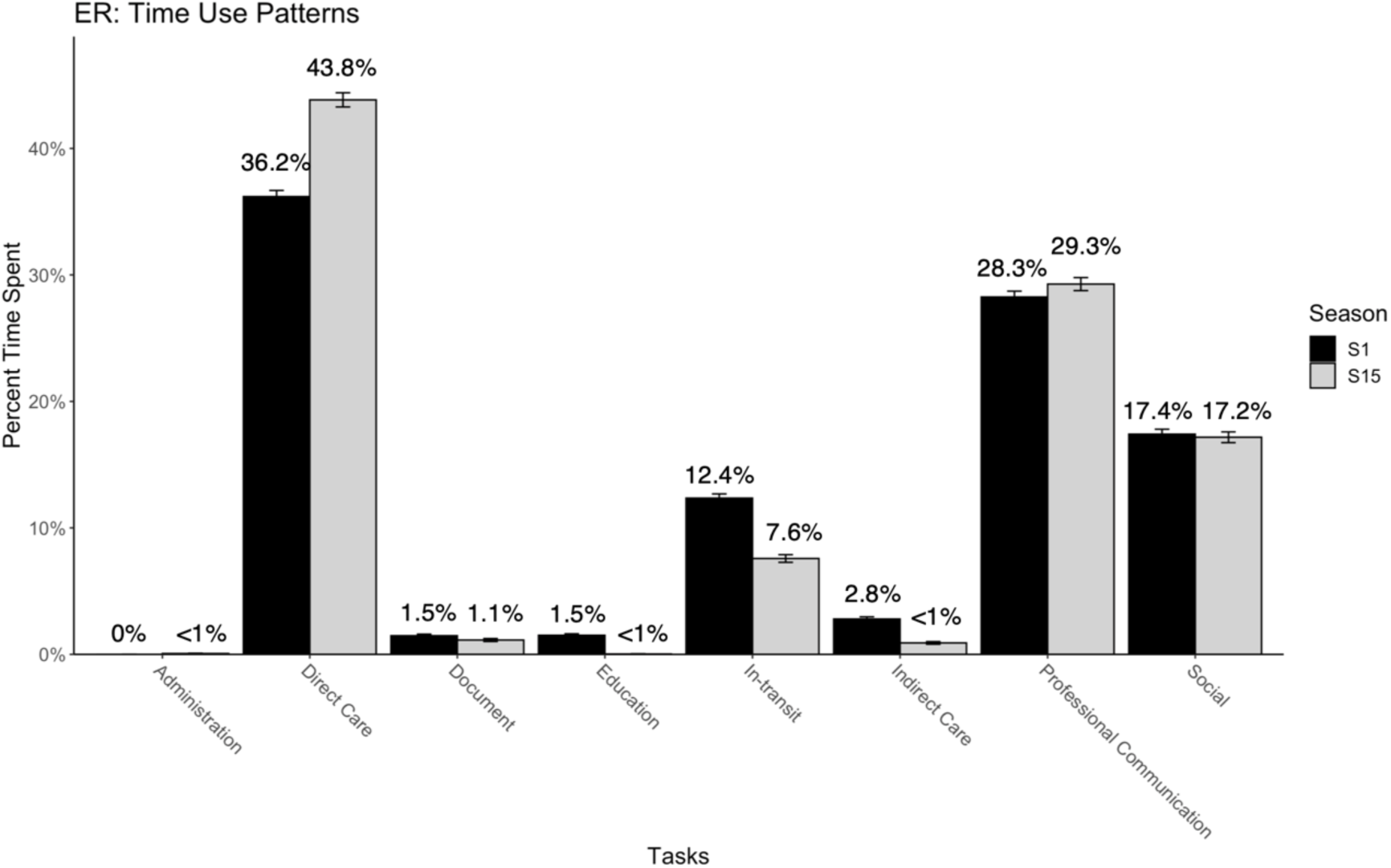
Percentage of time main characters spent on tasks during ER’s season 1 and season 15. Error bars are 95% confidence intervals.

**Figure 6.**
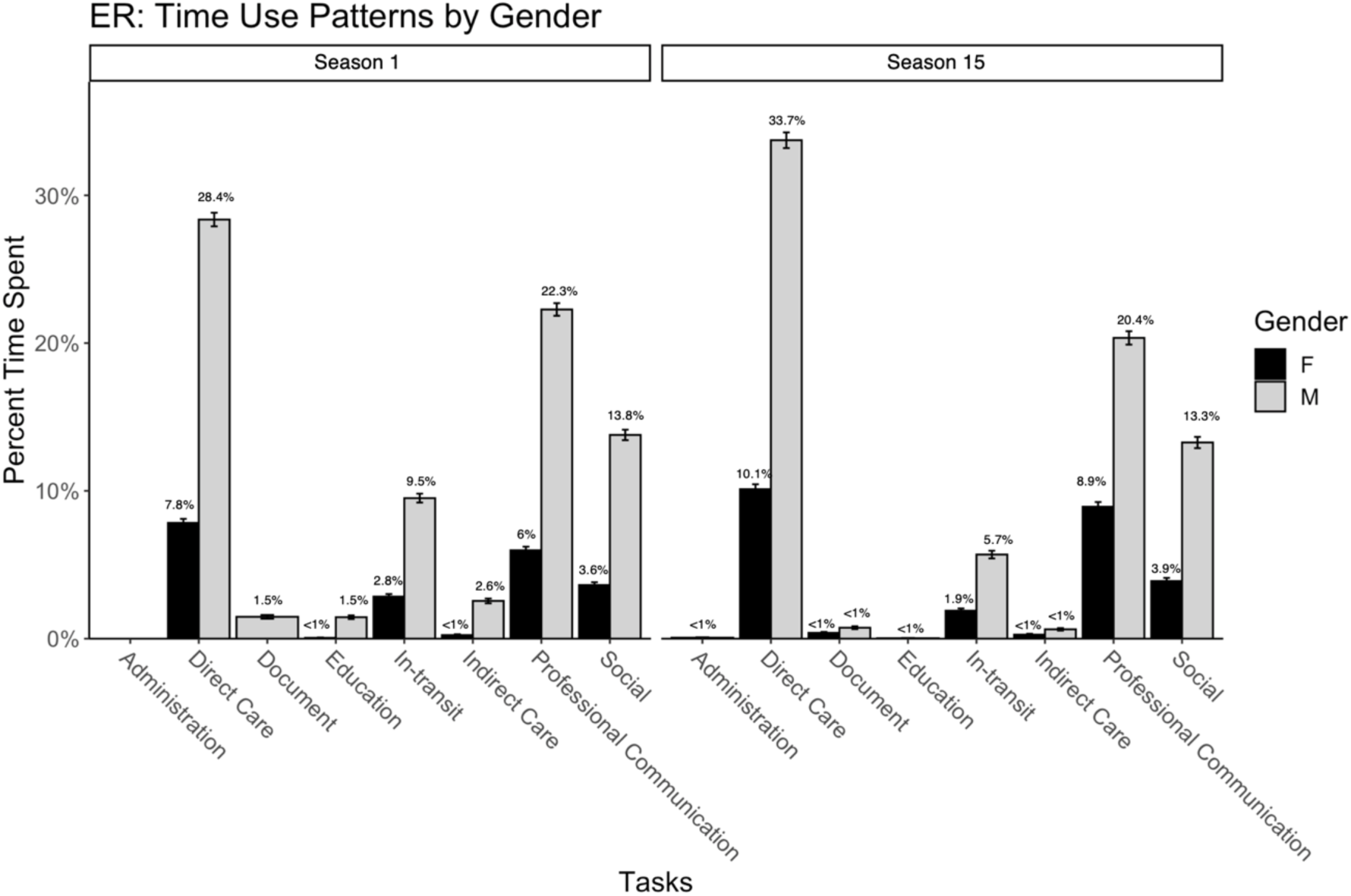
Percentage of time spent on tasks by gender during ER’s season 1 and season 15. Error bars are 95% confidence intervals.

A Fishers Exact test was performed to determine the relationship between ER seasons and the tasks performed. The relationship between these variables was significant, *p* < 0.001. On the other hand, we found a non-significant association between gender and time use patterns, *p* = 0.4762. Lastly, the ICC was computed to assess the agreement between two observers in observing physicians’ time use patterns in ER. Inter-rater reliability was good for ER, with an ICC of 0.89 and 0.81 for seasons 1 and 15, respectively.

## DISCUSSION

Overall, the medical dramas chosen do an adequate job of portraying the medical profession in media, apart from Grey’s Anatomy. Both seasons of Grey’s Anatomy dramatically overrepresent physicians’ time directly caring for patients and underrepresent their administrative duties compared to Ballerman et al. 2011, Westbrook et al. 2008, and Westbrook et al. 2010; but was close to Walter et al. 2014 in professional communication. Scrubs spent more time on social tasks than the other medical dramas, and when compared to other studies, Scrubs was similar to the daily duties of medical professionals, with direct care tasks (Season 1: 8.7%; Season 8: 3.5%) being reasonably close to Ballerman et al. 2011 (∼10%) [4], Göras et al. 2019 (4.4%) [18], and Walter et al. 2014 (8.2%) [17] but underrepresented when compared to Westbrook et al.2008 (15%) [7], and Westbrook et al. 2010 (28.6%) [16]. Additionally, Scrubs had similar time use patterns to Westbrook et al.2008 [7], Westbrook et al. 2010 [16], and Walter et al. 2014 [17] regarding professional communication but had lower activity patterns when compared to Ballerman et al. 2011 [4] and Göras et al. 2019 [18].

Comparatively, ER does well in capturing emergency medicine’s fast-paced and hectic nature. ER had the highest direct care tasks in season 1 at 36.2% and season 15 at 43.8% than the other medical dramas observed and the lowest in social tasks in season 1 at 17.4% and season 15 at 17.2%. Furthermore, direct care tasks were also observed to be either higher than or on par with other studies observing the emergency department [4, 7, 8, 16, 17]. Regardless of department, social tasks observed in hospital studies were either lower or on par with observations from ER [7, 8, 16, 17].

Our results showed that Grey’s Anatomy and Scrubs had statistically significant differences in clinician time use patterns by gender. Both medical shows favored one gender over the other. Females in Grey’s Anatomy dominated daily medical and social tasks for both seasons. Still, males increased their social time in season 16 while females decreased their direct care medical duties. On the other hand, Scrubs had a drastic difference in professional communication and social interactions between males and females, with males leading overall in both seasons. Additionally, both genders shared equal time spent on direct care tasks in season 8, but the activity only accounted for 1.9% (95%CI: 1.6% to 2.1%) for females and 1.7% (95%CI: 1.5% to 1.9%) for males which was a decrease for males in season 1 at 6.7% (95%CI: 6.4% to 7.0%). A description of the full nature of these differences – including whether these differences cast a positive or negative light on one gender is explainable by the relative prominence of actors or proportional to screen time, etc. is beyond the scope of this study.

Beyond the time use estimates themselves, this study does demonstrate consistently high observer agreement, as expressed in Intraclass Correlation Coefficients. Even the lowest ICC, seen in the last season of ER (0.81), would be considered “Good” under most circumstances. This suggests that, while the exact distribution of activities may not be relatable to actual clinical settings, this approach does provide a means of consistently and reliably familiarizing observers with the mechanics of recording observations and mapping what they see to broad categories of activity.

Medical dramas can serve as training tools when clinical observation is impossible. Medical dramas can be selected for their similarity to an in-person study. Beyond circumstances such as the COVID-19 pandemic, where observation is impossible, the use of medical dramas to train observers may also have use as an initial method of orienting and training, which can include avoiding or limiting patient care observations during this period. Additionally, using medical dramas with the WOMBAT software allows for a more flexible and accessible training period and not be limited to the HCP schedules. Direct observation of patient care is a time-consuming process for both the observer and the observed clinician and requires patients to consent for an observer to be present while they are being treated. Given these demands, a method for training observers on the software involved, familiarizing them with activity definitions, and addressing any questions that might arise that take place in a non-clinical setting may be extremely valuable.

One of the limitations of this study is that medical dramas do not accurately represent hospitals’ or healthcare professionals’ daily tasks. A direct comparison of the time use patterns from the selected medical dramas to real-world observations is unlikely to be directly productive, save for highlighting the differences in how the media portrays clinical medicine and the reality of it. Unsurprisingly, administrative duties and documentation were underrepresented in all medical dramas when compared to observations made in hospital observation studies [4, 7, 8, 16, 17]. This is not unreasonable – few would argue that charting and other administrative tasks make for riveting television.

For general medical settings, Grey’s Anatomy and Scrubs had excellent ICC scores, while ER had acceptably high ICC scores and might be more suitable for studies planned to involve the Emergency Department. Our methods reflect the ease of use and flexibility of medical dramas as a training tool for when in-person observations are limited or infeasible. When using these shows, however, one should be mindful that inaccuracies in the representation of clinical time use are present and that there are potentially biased portrayals of how male versus female clinicians spend their time. Given those limitations, however, we believe the use of medical dramas to train research staff in direct observation techniques is both a feasible and reliable technique.

### What is already known on this topic

- WOMBAT is a reliable tool for in-person hospital observation studies.
- Time use patterns of healthcare professionals have vast importance in understanding hospital burnout, contact patterns, and infectious disease spread.

### What this study adds

- Medical dramas can be used to train observers when in-person training is either impractical or infeasible.
- Our results have shown that observers trained on medical dramas have similar inter-rater reliability scores as those trained in a hospital setting.
- Medical dramas can be used as an initial training method to avoid or limit patient care observations.

## Data Availability

All data produced in the present study are available upon reasonable request to the authors.

## Appendix

